# Prescribing patterns for medical treatment of suspected prostatic obstruction: A spatiotemporal statistical analysis of Scottish open access data

**DOI:** 10.1101/2020.06.19.20135459

**Authors:** Federico Andreis, Richard Bryant, Emanuele Giorgi, Andrea Williamson, Ashleigh Ward

## Abstract

**Background:** Healthcare services treating men with prostate conditions are increasingly burdened worldwide. One of the competing factors in this demand is increasing diagnosis and treatment of lower urinary tract symptoms in men, much of which is suspected bladder outflow obstruction secondary to benign prostate hyperplasia/enlargement. However, the impact of increases on services is largely hidden, and there is limited knowledge of potential differences in management based on geography.

**Objective:** To investigate potential variation in the prescribing of drugs for suspected bladder outflow obstruction in Scotland based on analysis of publicly available data, and identify trends that may help to inform future prescribing behaviour.

**Design, setting, and participants:** We linked the relevant publicly available prescribing and patient data to all general practices in Scotland between October 2015 and November 2019.

**Outcome measurements and statistical analysis:** We analysed the numbers of daily doses of drugs prescribed for suspected bladder outflow obstruction per month using a Bayesian Poisson regression analysis, incorporating random effects to account for spatial and temporal elements in prescribing.

**Results:** Prescriptions of drugs to treat suspected bladder outflow obstruction increased during the observation period in Scotland, consistent with an ageing population and increased diagnosis. Whilst some determinants of health inequality regarding prescribing practices across health boards are consistent with those known from the literature, other inequalities remain unexplained after accounting for practice- and patient-specific characteristics such as socio-economic deprivation and rurality.

**Conclusions:** Variations in spatiotemporal prescribing for suspected bladder outflow obstruction exist in Scotland, some of which are unexplained and require further investigation.

## Introduction

The incidence of bothersome lower urinary tract symptoms (LUTS) in men, which are predominantly attributable to benign prostate hyperplasia (BPH) and/or benign prostatic enlargement (BPE) causing proven or suspected bladder outflow obstruction (BOO), is increasing globally [1] resulting in a reduced quality of life and increased healthcare expenditures [1, 2]. In Scotland as elsewhere in the United Kingdom and globally, some men may be managed conservatively or medically using drugs in the primary care setting for suspected or proven BOO (referred hereafter as BOO), whilst other men may have received investigation in a secondary care setting, before being recommended to receive medication in primary care. In Scotland healthcare services treating men with prostate conditions face an increasing burden from combinations of an ageing population, increased prostate cancer investigation and incidence [3], and increased awareness of male health issues. As a result of these combined factors, the impact of bothersome LUTS arising from BOO on service demand is largely hidden, and studies reporting their incidence and prevalence are predominantly cross-sectional in nature, with widely varying results [5].

Data regarding the prescription of drugs for the medical management of BOO as a cause for male bothersome LUTS has been used internationally in recent years to increase the understanding of patterns of prescribing within specific populations. This approach can be useful in order to identify potential demographic inequalities in patient’s access to this aspect of healthcare, and to understand the patient experience, predominantly using longitudinal datasets [6, 7, 8]. However, to date there is limited understanding of how the prescribing patterns for drugs used to medically treat BOO may differ both spatially and temporally. The increasing availability of high-quality open access healthcare data in Scotland has created the opportunity for a more refined and broader-based analysis of prescribing patterns for BOO. Moreover, Scotland’s diverse geography provides an ideal landscape to understand how factors that may impact on prescribing patterns differ spatially in terms of geography, as well as longitudinally [9]. We describe a statistical framework that allows the study of variations in the patterns of drug prescribing in both space and time, while accounting for numerous relevant socioeconomic and health-related factors. We have used this approach in order to statistically analyse publicly available data regarding the prescription of the two main classes of medications used to treat bothersome LUTS secondary to BOO in Scotland. Using this approach, we describe spatiotemporal trends that may help inform future urological practice for this common and ubiquitous condition.

## Material and Methods

### Data sources

Data was acquired and linked from numerous independent and publicly accessible sources. The dataset comprised information at the level of individual General Practices (GPs) and was been created from the following four sources:

1. NHS Scotland OpenData: drug prescriptions monthly data (https://www.opendata.nhs.scot/dataset/prescriptions-in-the-community)
2. Information Services Division Scotland (ISD): practice details, deprivation, rurality (https://www.isdscotland.org/Health-Topics/General-Practice/)
3. National Institute for Health and Care Excellence (NICE): daily dosages (https://www.nice.org.uk)
4. National Records of Scotland (NRS): postcodes, health boards (https://www.nrscotland.gov.uk/statistics-and-data/geography/our-products/scottish-postcode-directory/2018-2)

The keys used to link the sources were the unique GP code and date. The ISD and NRS portals provided information regarding both the medical practice (i.e. GP) and their patient population. Table 1 contains a list of the variables extracted from these sources. We used both socio-economic deprivation and rurality to account for socio-demographic status. In particular, we used the 2016 Scottish Index of Multiple Deprivation (SIMD) [10] and a recoded version of the 2018 Scottish Government Urban Rural Classification [11]. Additionally, the ISD and NRS sources provided GP-level information regarding the size of the individual practice patient list, the gender and age distribution, and the location of the practice (based on postcode and name of relevant Health Board). It is important to note that digital boundaries of Scottish GP practice catchment areas are not available; as a consequence, the spatial reference for both the GP practice- and patient-level data is the GP practice address. All GP practices open for the whole study period (October 2015 to November 2019) were included in the dataset (903 of 944, 95.7%, practices in Scotland open as of 1^st^ January 2019). The Scottish Open Data portal provided the prescribed amounts of the drugs of interest per month.

**Table 1.** Variables extracted from the four data sources and used as covariates in the statistical model.

| Variable | Type | Short description |
| --- | --- | --- |
| Drug group | Categorical | α-1 blocker or 5-α reductase inhibitor |
| GP patient list size | Numerical, time-varying | Number of registered patients per GP practice |
| GP practice run | Categorical, baseline | Is the practice run by GPs (contract type 17J and 17C), rather than by the NHS (contract type 2C)? Yes/No |
| Dispensing GP practice | Categorical, baseline | Does the GP practice have a license to dispense medicines? Yes/No |
| Males 45p | Numerical, time-varying | Proportion of patients that are males aged $\geq 45$ years |
| Deprived 15 | Numerical, time-varying | Proportion of patients under the 15<sup>th</sup> national percentile of deprivation (SIMD) |
| Remote | Categorical, time-varying | Modal rurality index among the patients (Scottish Government 8-fold Urban Rural Classification) recoded as remote (4, 5, 7, 8) or non-remote (1, 2, 3, 6). Yes/No |
| Postcode | Categorical, baseline | GP practice postcode as of 2019 |
| Health board | Categorical, baseline | Name of Scottish Health Board where the GP practice is situated. |

### BOO medications analysed

We specifically focussed on the prescribing of α−1 blocking drugs and 5−α reductase inhibitors as these are the only specific medications used for BOO. α−1 blockers are the first-line and most commonly used drugs prescribed for LUTS secondary to BOO [1, 12], whilst 5−α reductase inhibitors are generally recommended where men are considered at high risk of BPH progression due to a significantly enlarged prostate, either as monotherapy or in combination with α−1 blockers [1, 12, 13]. The α−1 blockers alfuzosin and tamsulosin hydrochloride were included in the study, however doxazosin and terazosin were not included as the amounts of these drugs prescribed were negligible, whilst doxazosin may be prescribed for hypertension. The 5−α reductase inhibitors dutasteride and finasteride were both included in this study. When combined with the suggested daily dosages from the NICE website (see Table 2), a proxy for the number of daily doses was constructed by rounding to the nearest integer the following expression:

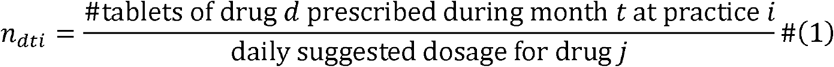

**Table 2.**
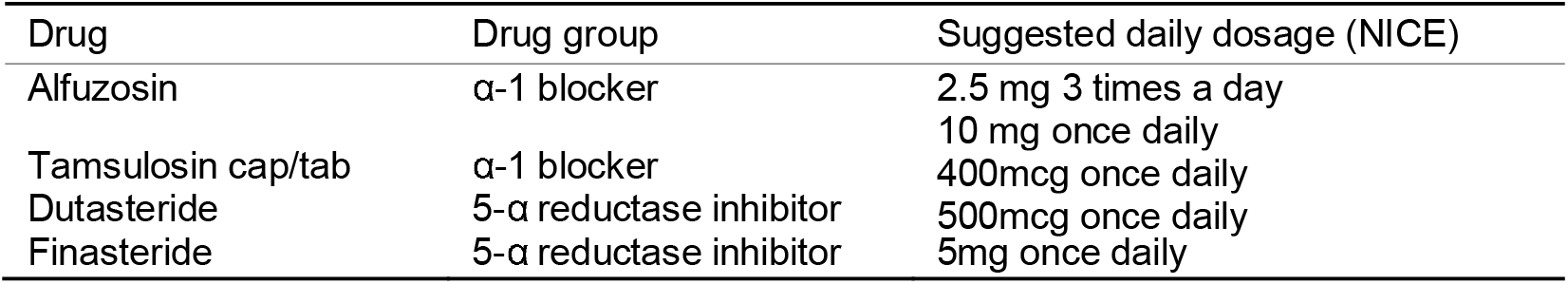
Suggested daily dosages of the study α-1 blockers and 5-α reductase inhibitors according to NICE guidelines.

where *i, d*, and *t* represent the GP practice, drug, and month respectively. In our dataset, we have observations for *i* = 1, …,903 GP practices over *t*= 1, …, 50 months; *d* indexes the drugs reported in Table 2. The number of daily doses for each of the drug groups was then constructed by summation of the respective drugs’ monthly amounts of daily doses. We denote with *n*^*jti*^ the number of daily doses of drugs belonging to group *j*∈ {α1 − blocker, 5 − α reductase inhibitor}, prescribed during month *t*, at each GP practice *i*.

We then used a Poisson model in order to describe the average number of daily doses of BOO medication prescribed each month, accounting for each of the covariates presented in Table 1, with additional spatial and temporal random effects. Possible interactions between the prescribing pattern of individual GP practices, or their geographical location, with the prescribed drug type were also investigated. Equation 1 describes the model’s functional form, while Table 3 summarises our choices in terms of spatial and temporal structure. Further details of the statistical analysis are provided as supplementary material.

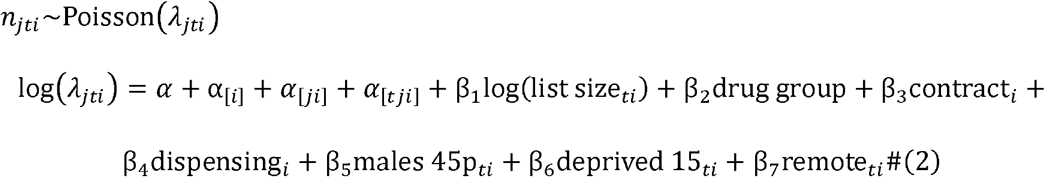

where *j* (drug group), *t* (month), and *i* (each GP practice) are as described, and *α*_[i]_, *α* _[ji]_, *α* _[tji]_ denote the random effects, summarised in Table 3. A detailed discussion of the choice of the random components of the model is available in the technical supplement.

**Table 3.** Spatial and temporal random effects.

| Parameter | Type | Short description |
| --- | --- | --- |
| $\alpha_{[i]}$ | Spatial | intrinsic Conditional Auto Regressive (iCAR) |
| $\alpha_{[ji]}$ | Unstructured | Unstructured interaction between GP and drug type |
| $\alpha_{[tji]}$ | Temporal | First-order Auto Regressive (AR1), grouped by health board and drug type |

### Computational analysis

All computational analyses were carried out in R 3.6.1 [14]. We estimated the model parameters within the Integrated Nested Laplace Approximation framework using the inla package [15].

## Results

We investigated the effects of each the available covariates (including GP patient list size; BOO drug group; whether the practice was GP run; whether the GP practice dispensed medication; proportion of males aged 45 years; higher proportion of socio-economic deprivation; and remote/rural location) on the study outcome. Table 4 summarises the output of the Poisson regression estimates of each of the associated parameters. In a further analysis, we interpreted each of these parameters in terms of relative differences from the model average (with the exception of GP practice list size).

**Table 4.** Summary of estimated posterior distributions for fixed effects. This summarises the model estimates of the fixed effect, and the uncertainty surrounding the estimates.

|  | Mean | SD | 2.5% | 50% | 97.5% | Mode |
| --- | --- | --- | --- | --- | --- | --- |
| $\alpha$ – Intercept | -1.570 | 0.040 | -1.648 | -1.570 | -1.492 | -1.570 |
| $\beta_1$ – log(GP patient list size) | 0.915 | 0.002 | 0.911 | 0.915 | 0.919 | 0.915 |
| $\beta_2$ – Drug group (5-α reductase inhibitors) | -0.625 | 0.017 | -0.658 | -0.625 | -0.591 | -0.625 |
| $\beta_3$ – GP run practices | 0.238 | 0.033 | 0.174 | 0.238 | 0.302 | 0.238 |
| $\beta_4$ – Dispensing GP practice | -0.086 | 0.026 | -0.138 | -0.086 | -0.035 | -0.086 |
| $\beta_5$ – Males aged $\geq 45$ years | 4.816 | 0.032 | 4.754 | 4.816 | 4.879 | 4.816 |
| $\beta_6$ – Socially deprived area | 0.017 | 0.007 | 0.003 | 0.017 | 0.030 | 0.017 |
| $\beta_7$ – Remote/rural GP practice | 0.002 | >0.001 | 0.002 | 0.002 | 0.003 | 0.002 |

The GP practice patient list size was observed to have an effect consistent with an estimated increase of slightly less than one daily dose per month per additional patient (point estimate 0.915). As expected, the prescribed doses of the 5−α reductase inhibitor drug group were observed to be on average less than the prescribed doses of the α − 1 blocker drug group. The estimated relative difference was calculated to be on average 45-48% fewer 5 − α reductase inhibitor drug prescriptions than α − 1 blocker drug prescriptions per month (point estimate 46%).

We observed that the nature of how the GP practices are individually run are associated with BOO drug-prescribing practice, with GP-run practices (rather than by direct Health Board-run practices) having larger volumes of these prescriptions. The model results are consistent with a 19-35% (point estimate 27%) increased number of monthly prescriptions for GP-run practices compared with Health Board-run practices, all else being equal. This is an interesting result, and to the best of our knowledge no rigorous research has been undertaken to investigate the role that how a GP practice is managed plays on the stability of GP provision of care and, in turn, on patient help seeking behaviours and clinical outcomes. Conversely, we observed that the small number of GP practices who run their own dispensary are associated with 3-13% (point estimate 8%) lower volumes of BOO drug prescriptions, compared to GP practices who do not, keeping all other variables constant. Practices that dispense tend to do so because there is no local community pharmacy, and one is needed for patient care. It is plausible that having a dispensing licence may reflect an aspect of remoteness that is not captured by the rurality measure we employ. We found the proportion of males aged ≥45 years in the GP practice patient list to be positively associated with the volume of BOO prescriptions, with an estimated average increase of 4.9% monthly prescriptions per percentage point. In contrast, the levels of socio-economic deprivation, and a remote/rural geographical location of each GP practice, were found to have a negligible association with prescribing practice.

Next, we investigated the statistical model’s interpolations according to Health Board and drug group (α−1 blocking drugs and 5−α reductase inhibitors) (**Figure 1**). This analysis highlighted a general increase in the volumes of prescriptions for these medications over the 4-year study period. Trends in prescribing were observed to be broadly similar across most Health Boards, with a less prominent increase in less populated areas, which may potentially be attributable to a more stable population structure over the 4-year period of observation, as well as unaccounted for differences in help seeking behaviours. Within most Health Boards the growth in prescriptions for each drug group were almost parallel during the four-year observation period, suggesting consistency in terms of increased BOO drug prescribing. Additional observations included a marked shift in volumes of α−1 blocking drug prescriptions in Lanarkshire between November 2016 and February 2017, however no further specific information regarding this phenomenon was available within the publicly available dataset. In the absence of a reasonable explanation for this singular observation in this one Health Board, this has been treated as an artefact due to a change in data-recording.

**Figure 1:**
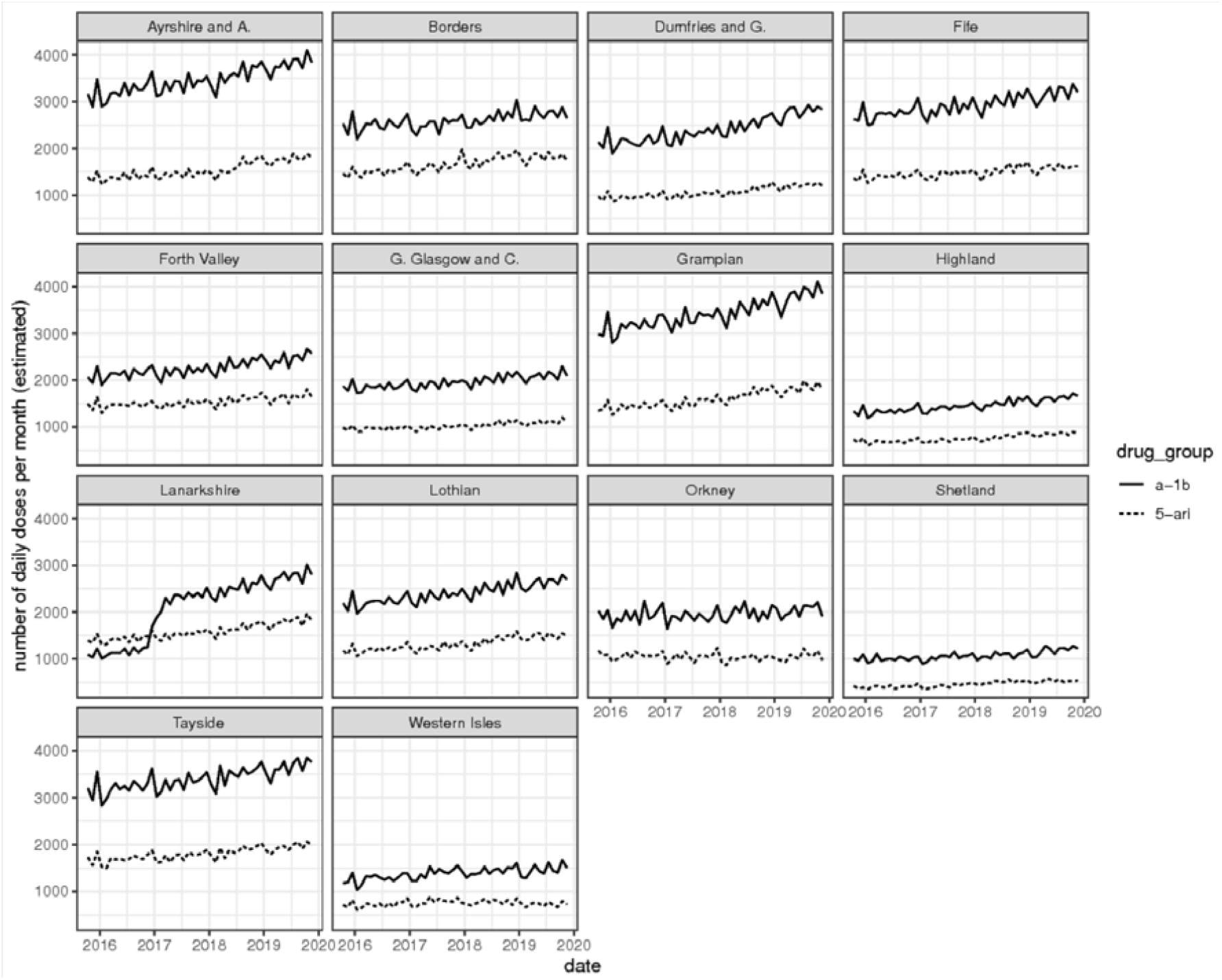
Statistical model average number of daily doses of each BOO drug type prescribed per month, by individual Scottish Health Board and adjusted for GP practice patient roll size.

Next, we investigated how the statistical model residuals may provide insight into how BOO drug prescribing behaviour differed across Scotland during the 4-year study period. **Figure 2** illustrates histograms of exponentiated residuals for each individual Health Board across Scotland, where a reference value of 1= *e*^0^ represents a null residual (i.e. there is no deviation from the model’s predicted average). Each residual is specific to one individual anonymous GP practice. On an exponential scale, a value of 1 indicates a prescribing behaviour in line with the average as described by the statistical model across Scotland during the study period. A value below 1 indicates prescribing volumes higher than expected based on the model, whilst a value larger than 1 indicates prescribing volumes lower than expected. Asymmetry around the reference value of 1, and/or multiple modes of distribution, suggests a different behaviour of individual GP practices within a Health Board compared against the national average across Scotland.

**Figure 2:**
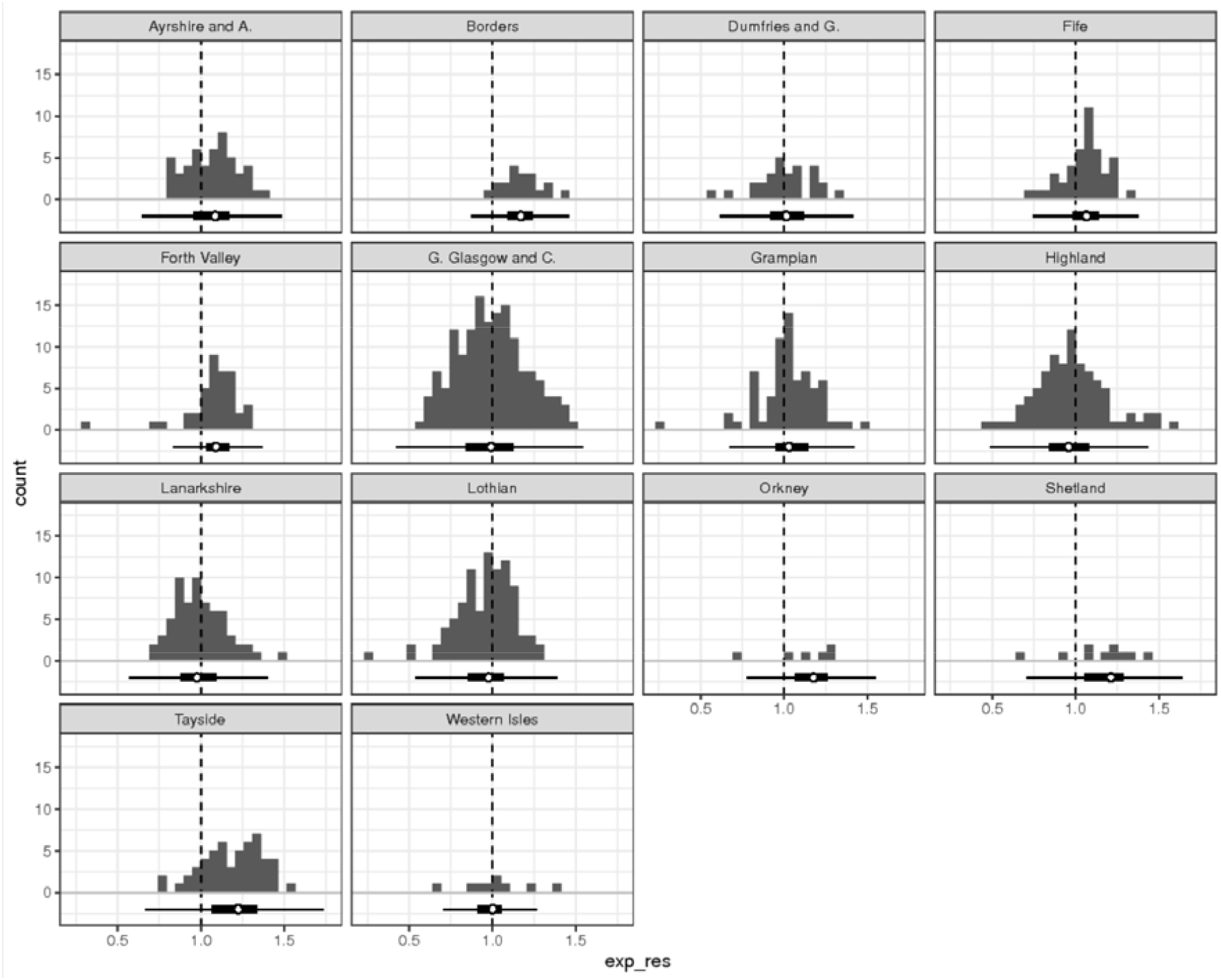
Frequency histograms of the statistical model’s exponentiated residuals for each Scottish Health Board. The vertical dashed line represents the null residual (value=1 on the exponentiated scale), and each small black square represents the average number of daily prescriptions of BOO drugs (combined α − 1 blockers and 5−α reductase inhibitors) per month per individual anonymous GP practice within each Health Board. The boxplot below each histogram illustrates the same data distribution (the white point marks the median, the thick line represents the usual box, and the thin line represents the whiskers). Ideally the exponentiated residuals for each GP practice would lie around the null residual (i.e. value=1), however the heterogeneity in prescribing practice is illustrated by increased spread to the left and right of the null residual. A shift of GP practice distributions to the right of the null residual, as seen in Health Boards such as Borders, Fife, Orkney, Shetland and Tayside, illustrates less prescribing of these medications than would be expected.

Using this statistical approach, we generally observed consistency in the prescription of drugs for BOO across Health Boards in Scotland, with most distributions appearing to be approximately unimodal and symmetric around the null residual (i.e. the reference value of 1= *e*^0^). However, some interesting patterns of prescribing can be observed, such as the reduced level of BOO drug prescribing in some individual Health Boards (most notably Borders, Forth Valley, Tayside, Orkney and Shetland) compared to the average prescribing rate across Scotland. The reasons for these differences in prescribing pattern in these individual Health Boards are unclear and require further investigation.

Next, we examined the BOO drug prescribing patterns of GP practices with the aim of assessing and visualising possible spatial patterns. Once again using the model residuals, it was possible to identify individual anonymous practices at the extremes of the BOO prescribing distribution, and show their Health Board location on a map. This approach can be useful in order to detect geographic clusters of similar prescribing patterns, while accounting for temporal trends and other potential confounding factors that are already included in the model. For example, the policymaker might be interested in identifying clusters of GP practices where the rates of prescribing are either very low or very high with respect to the national average. We present the results of such analysis in **Figure 3** where, to preserve anonymity of the individual practices, we have aggregated the information at the Health Board level. We denote those GP practices with a model residual below the 2.5% percentile of the overall distribution of residuals as “high-volumes prescribers”, and those above the 97.5% percentile as “low-volumes prescribers”. We stress that high- and low- are with respect to the national average as described by the model Moreover, the quantile thresholds are arbitrary and can be adjusted according to the analytical need.

**Figure 3:**
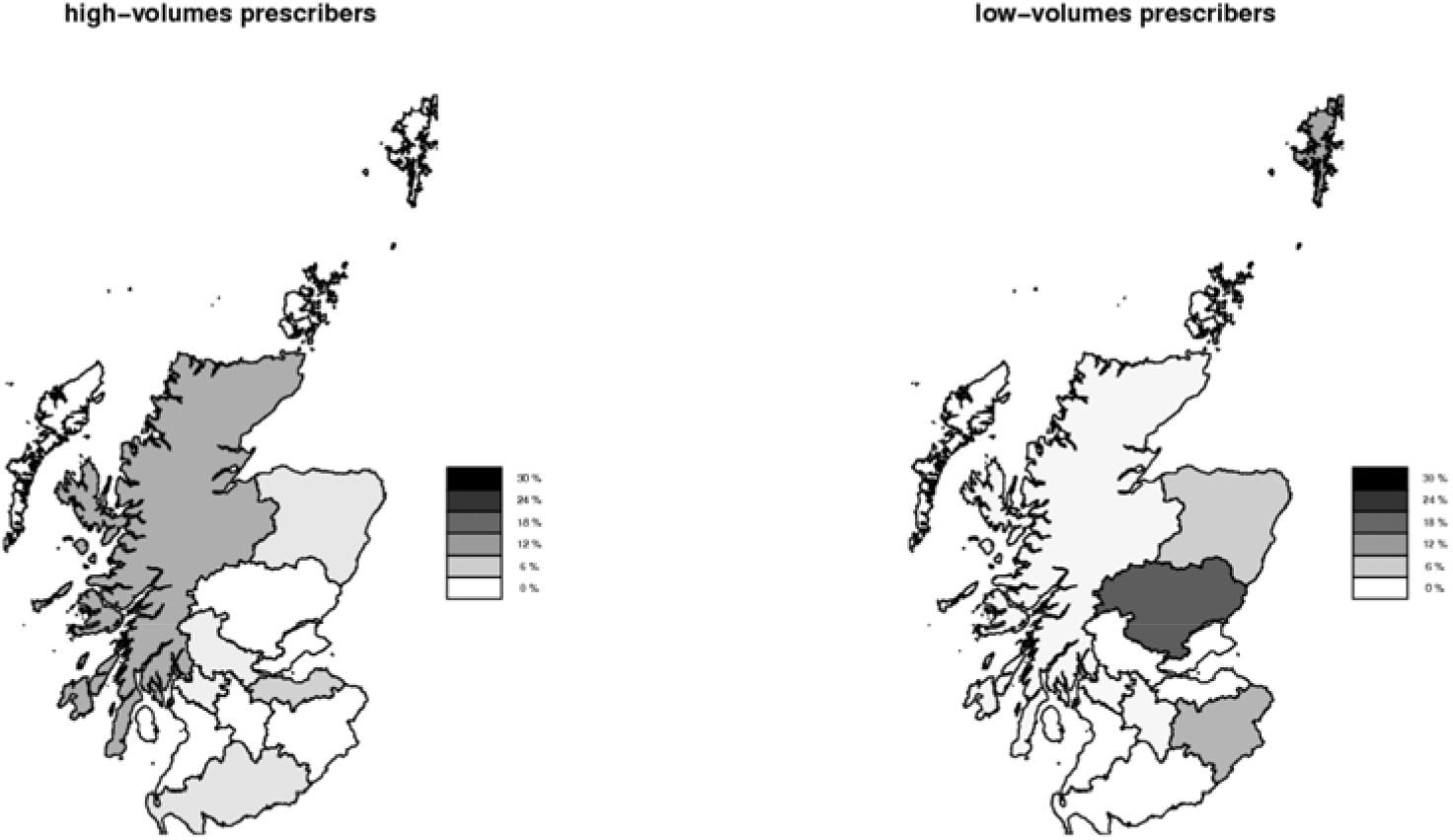
Maps of Scotland highlighting the Health Board locations of anonymous GP practices at the extremes of the distribution of BOO drug prescribing. The left panel shows the percentage of GP practices within each Health Board that have been identified (via model residuals) as being high in prescribing volumes with respect to the national average (i.e. model residuals <2.5% percentile). Similarly, the right panel shows the percentage of GP practices low in prescribing volumes with respect to the national average (i.e. model residuals >97.5% percentile).

We observed that Highland is the Health Board with the largest percentage (∼9.5%) of GP practices that were identified as being in the upper tail of prescribing volumes (identified by model residuals <2.5% percentile), followed by Lothian (∼5.3%), Dumfries and Galloway (∼3.2%), Grampian (∼2.9%), Forth Valley (∼2%) and Greater Glasgow and Clyde (∼1.7%). No high-volume prescribing GP practices were found within the remaining Health Boards. On the other hand, Tayside was identified as the Health Board with the largest percentage (∼19%) of low-volume prescribers (identified by model residuals >97.5% percentile). Shetland (10%), Borders (∼8.7%), Grampian (∼5.7%), Highland (∼1.1%), Lothian (∼1%), and Greater Glasgow and Clyde (∼0.9%) also contained GP practices that prescribed far below the national average. The remaining Health Boards did not contain any low-volume prescribing practices. The underlying reasons for these observations are currently unclear and require further investigation.

## Discussion

This study has investigated patterns of medical prescribing for the two most common classes of drug (i.e. α − 1 blockers and 5−α reductase inhibitors) used to treat BOO in Scotland over a recent 4-year period (October 2015 - November 2019) using publicly available data from individual GP practices across all Health Boards. Using this approach we generated a study dataset by linking prescribing data to GP practice-specific information (such as their type of contract with NHS, and their licence to dispense) and summary characteristics of their patient populations (such as patient age, and GP practice-associated levels of socio-economic deprivation and rurality). This has enabled us to develop an improved understanding of patterns of prescribing of these medications across Scotland, and identify where prescription volumes may vary, having accounted for these variables.

A trend of increased BOO drug prescribing practice was observed throughout the 4-year observation window consistent with an ageing Scottish population, and perhaps also due to an increased awareness of male health issues amongst both patients and general practice teams, and increased referral of men to secondary care due to a raised PSA, which may indicate BPH as well as possible prostate cancer [16, 17]. However, it would be interesting to understand more fully the reasons behind the observed increase in prescribing across all Scottish Health Boards, and the role that increased awareness and patient help seeking behaviour may play in this phenomenon. It may also be interesting to identify the relative contribution (or otherwise) made by increased referral to secondary care with suspected prostate cancer (as identified by a raised PSA), which might result in increased BOO/BPH prescribing as a secondary consequence of a raised PSA referral. A further possibility may be that an increased focus on cancer management, rather than BPH surgery, may lead to an increase in medical drug use for BOO, rather than definitive intervention – this is speculative, but would be very interesting to investigate in future studies, particularly in order to investigate how this may vary geographically. Although it has been acknowledged that aspects regarding individual GP medical practices may potentially contribute to prostate health inequalities [18, 19], to our knowledge this is the first time that the particular characteristics of a GP practice (such as patient ages, GP-run practices, dispensing ability, socio-economic deprivation levels, and rurality) have been identified as being potential factors influencing BOO drug prescribing patterns. It was interesting to observe that whether a practice was Health Board or GP-run, and whether a dispensing pharmacy was present within the GP practice, was associated with higher and lower prescribing volumes respectively. While the individual characteristics of GP medical practices are known to differ by region [20], research accounting for these factors is relatively sparse; hence it is unclear why this difference exists.

This study has several limitations. Firstly, the available data only exists in the public domain at an aggregate (i.e. individual GP practice or patient list) level, and further information would be helpful in order to identify the reasons why prescribing practices vary at a more granular level (perhaps by associating the prescribing practice with the presence or absence of a GP with a particular interest in men’s health issues, as an example). Secondly, the use of summary data regarding levels of socio-economic deprivation and rurality of patients reduces the accuracy with which one can estimate the shape of their association with outcome, with these factors likely to be important in terms of access to healthcare and prescribing. Similarly, not having information about specific GP practice catchment areas made it necessary to use an adjacency matrix based on postcodes, rather than actual distances, resulting in a sub-optimal method of accounting for underlying spatial processes; furthermore, future work utilising anonymised individual-level patient data would also help to increase accuracy. Finally, it would be useful to analyse potential relationships between BOO drug prescribing patterns and longer-term outcomes from BOO in terms of clinical progression of this condition, such as need for future surgical intervention or treatment for BOO complications, such as acute urinary retention, and how they relate to investigation for suspected cancer or raised PSA. Such an analysis requires additional hospital data which in future work could be accessed via established high-quality data linkage systems in the Scottish National Safehaven.

It would also be interesting to understand the relative contribution of patient ageing to the rate of rise in BOO drugs prescription, and the degree to which other factors such as help-seeking behaviour and clinician awareness might be contributing to this rate of rise in prescription volumes. These possibilities require further investigation. Nevertheless, despite these caveats and limitations, the analyses presented herein have identified interesting BOO medical prescribing trends across Scotland that require further investigation in future studies.

In addition to identifying influences from spatial, temporal, and socioeconomic factors in regards to volumes of prescription of BOO medications, our analysis suggests that other sources of variability might account for inequalities in prescribing practice. In recent decades a considerable amount of research has been undertaken in order to better understand complex health-related phenomena whose determinants might transcend socio-demographic factors, and can best be described by a unique regional culture or a strong ‘social patterning’ [21, 22]. Although at this stage it is unclear what the remaining hidden determinants of the variations in BOO drug-prescribing patterns may be, Scotland’s cultural diversity as described in the wider literature may constitute a potential explanation [9]. This warrants further investigation in future research.

It would be interesting for future research to investigate trends and potential inequalities regarding all prostate-related conditions (both benign and malignant) over space and time in other settings and geographical locations. In particular, it would be interesting to investigate whether the trends observed in Scotland may similarly exist in other regions of the UK and Ireland, and if so, are variations attributable to common factors such as remoteness/rurality and socio-economic status. Further research to investigate medical practice phenomena as complex as prescribing patterns using publicly available data may identify key limitations in practice, and may inform data collection and data sharing in the future. In addition, the general nature of the statistical modelling framework we have proposed in this study may be extended to investigate other kinds of models, covariates, and medical conditions in a range of future studies.

## Conclusions

The volume of prescriptions of drugs for LUTS secondary to BOO has steadily increased across Scotland during a recent 4-year observation period, which may, at least in part, be attributable to an ageing population and increased diagnoses. Prescription volume for these medications is associated with various characteristics of the GP practice (such as type of contract and presence of a licence to dispense). However, some inequalities in prescribing across Health Boards in Scotland remain unexplained after accounting for GP practice- and patient-specific characteristics such as socio-economic deprivation and rurality, suggesting there may be other hitherto unknown determinants of BOO drug prescribing practice. Further research is warranted to understand these differences more fully.

## Data Availability

All data referred to in the manuscript is freely and openly available online through the sources therein indicated. Moreover, the dataset and shapefiles used are provided as supplementary material.

https://www.dropbox.com/s/i5h0r8lv0jyqmcd/Data.zip?dl=0

## Acknowledgements

The authors would like to acknowledge support and encouragement received by Prof Alan McNeill (NHS Scotland) and Prostate Scotland at the initial conception of this project.

